# Don’t stop the heart: a performance analysis of large language models and potassium dosing

**DOI:** 10.64898/2026.06.02.26354762

**Authors:** Kaitlin Blotske, Xingmeng Zhao, Kelli Henry, Brian Murray, Yanjun Gao, Susan E. Smith, Nathaniel Wayne, Pam Ku, Brooke Smith, Stacy Moua, Andrea Sikora

**Author notes:** **Funding:** Funding through Agency of Healthcare Research and Quality for Dr. Sikora was provided through R21HS028485 and R01HS029009. Funding through National Library of Medicine for Dr. Gao was provided through R00LM014308.

## Abstract

**Background:** Electrolyte replacement is ubiquitous in the acute care setting, but its familiarity cannot belie that even small dosing errors with potassium can cause lethal cardiac arrhythmias. Recently, MedAgentBench offered a benchmark for agentic artificial intelligence (AI) including the ability to correctly dose potassium based on a single rule; however, this does not adequately reflect the clinical complexity or safety concerns of an agent that has been used as the lethal injection. The purpose of this analysis was to a probe leaderboard large language model (LLM) capabilities to follow basic dosing rules to safely replace potassium in a series of clinician-annotated cases.

**Methods:** Using a clinician panel, we developed a series of dosing principles and 20 clinical cases reflective of the complexity of potassium replacement. External clinicians were surveyed to assess practice variability and agreement to clinician panel answers. We tested GPT-5-chat with each case in triplicate, with and without the clinician curated dosing principles, and prompted the model to answer six questions involving potassium goals, dosing, route, lab frequency, concurrent interventions, and the model’s perceived level of confidence for the output and complexity of the case. The primary outcome was the rate of appropriate recommendations in comparison to clinician answers.

**Results:** A total of 54 clinicians reviewed the 20 hypokalemia cases and hypokalemia dosing guideline. Clinicians expressed “highly agree” or “somewhat agree” for 66.8% of the cases evaluated when asked if they agree with the guideline-recommended management. When given the potassium dosing guideline, total errors dropped from 165 to 104, and average accuracy improved from 45% to 65% with GPT-5-Chat. GPT-5-Chat conveyed a high level of confidence for 100% of responses, while labeling 80% and 76% of cases as highly complex with and without the criteria, respectively. Potential harm scores were considerable in both groups, however, a notable reduction in severity scores occurred with the dosing guidance document. Recommendations on concurrent interventions and dosing had the highest rate of errors in both groups.

**Conclusions:** Benchmarks must appropriately reflect clinical complexity to be considered valuable for the deployment of agentic artificial intelligence tools in the healthcare domain. GPT-5-Chat assessment on a comprehensive medication management task for potassium replacement showed improvement with dosing guidance, yet unfit benchmarking performance.

## Introduction

Worldwide, an estimated 1 in 30 patients likely to experience some form of medication-related harm, with 50% being preventable medication errors.^1^ In 2021, two pharmacists caught 634 medication errors in just six weeks at an academic medical center; this finding is in line with a myriad of prior reports.^2^ Furthermore, roughly 25% of medication error events are expected to result in severe or life-threatening patient harm.^1^ To ensure safe handling of medications and tracking of medication errors, the Institute of Safe Medication Practice (ISMP) was developed.^3^ This organization publishes guidelines and tools to reduce the risk of medication related events occurring across healthcare settings. One tool, the ISMP High-Alert Medication list, comprises drugs that have high potential for significant patient harm.^4^ One of these high-risk medications, potassium, is an essential electrolyte for normal functioning of all cells and regulation of action potentials, notably in the heart.^5^

Electrolyte replacement is one of the most common tasks occurring in acute care settings and is often more complex, than a face-value lab result suggests. In the case of potassium replacement, various factors must be considered to optimize and ensure safe administration (i.e., renal function or dialysis, comorbidities or acute problems, other electrolytes, medications, route, type of formulation, administration rate, etc.). In current practice, institutions often replenish potassium through modalities of provider- or nurse/pharmacist-driven protocols or order sets. Though these practices are fixed and inflexible in the highly dynamic environment of a patient’s clinical course.

Recently, MedAgentBench published a virtual electronic health record (EHR) benchmark stating their system allowed for successful benchmarking of various large language models to complete tasks related to laboratory test result, vital sign, procedural order, diagnosis, and medication ordering.^6^ For one medication ordering task, this model employed a single rule for potassium dosing where under a serum level threshold of 3.5 mEq/L the model was instructed to dose “10 mEq of oral potassium for every 0.1 mEq/L below the specified threshold.” This rule, although used at times in practice, does not adequately reflect clinical complexities and risks severe or life-threatening harm in specific scenarios (i.e. diabetic ketoacidosis, refeeding syndrome, etc.).

Leveraging artificial intelligence to optimize non-linear clinical settings is an attractive addition in practice for the purpose of reducing workload burden and mental strain. However, in order to know if an agent could effectively function in a clinical space, rigorous pre-clinical and clinical testing are essential to ensure the safety of this approach. The purpose of this evaluation is to test a novel large language model, GPT-5-Chat, on complex potassium rules and criteria using hypokalemia patient vignettes to assess safe potassium dosing performance.

## Methods

### Study Design

The purpose of this study was to generate a series of clinician-annotated and complex hypokalemia cases to evaluate the capabilities of a leaderboard LLM to follow curated dosing rules to safely replace potassium.

This project was reviewed and approved by the University of Colorado Institutional Review Board (COMIRB #24-2170). All methods were performed in accordance with the ethical standards of the Helsinki Declaration of 1975.^7^ This evaluation will follow the transparent reporting of a multivariable model for individual prognosis or diagnosis (TRIPOD–LLM) extension reporting frameworks.^8^

### Potassium Dosing Criterion and Rules

To capture the multifaceted considerations for safe and effective replenishment of potassium, a clinician panel of seven pharmacists, curated a guidance document containing special situations that affect potassium dosing rules, frequency of lab monitoring, serum potassium goals, and additional interventions relating to hypokalemia. The guidance document also comprised a list of negotiable and non-negotiable (i.e. do NOT administer undiluted or intravenous push) rules as safety thresholds for potassium administration **(Supplemental Appendix)**. The purpose of this guidance document was to create a comprehensive tool for approaching clinical scenarios with hypokalemic patients.

### Potassium Cases Dataset

A dataset including 20 hypokalemic patient vignettes was created by two clinical pharmacists. Each case contained a short patient case description, pertinent laboratory results, vital signs, and repeat lab order frequency. Cases varied in levels of complexity (i.e. acute kidney injury, diabetic ketoacidosis, refeeding syndrome, etc.), containing one or more clinical scenarios that can affect a clinician’s approach to potassium dosing recommendations. A clinician panel of 7 pharmacists reviewed and agreed upon answers for each case based on the potassium dosing guidance document. The dataset was developed to assess large language model performance in providing safe and accurate recommendations for treating hypokalemic patients.

### Clinician Survey

A survey containing all 20 patient cases and the curated potassium dosing rules was created for the purpose of assessing practice variability within potassium dosing. Survey was distributed to three academic medical centers, UCHealth University of Colorado Hospital (UCH), Wellstar Medical College of Georgia (MCG) Health Medical Center, and the University of North Carolina (UNC). Criteria for participation included the following clinician types: physician (PGY2 residents or above), pharmacists (PGY2 residents or above), physician assistants, and nurse practitioners. Baseline characteristics collected from respondents were site of practice, clinician role, specialty or training area, patient level of care/acuity, highest level of training, highest formal degree, and years of experience outside of training.

Patient case questions evaluated clinician preference for potassium goal, dose, route of administration, frequency of labs, additional interventions. Providers were also asked to categorize overall complexity of the patient case and their confidence for answer choices. After initial assessment, clinicians were provided the same patient case with investigator team answers to the case from the curated potassium dosing rules. Clinicians were asked to rate their level of agreement with the investigator teams responses, their level of confidence in answering cases, and the level of complexity for each case. The survey was developed and administered via the RedCap^®^ platform.^9^

### Preceptor-student recordings

Virtual sessions of student assessment and recommendations with preceptor oversight of hypokalemia cases were conducted to capture pharmacist preceptor and student dialogue within a learning environment. Preceptor and pharmacy candidate participants were selected via volunteers from university campuses with School of Pharmacy. Each student was presented with one of the potassium vignettes comprising a short description and laboratory values. Pharmacy preceptor volunteers were given the same case with answers to help guide discussion and student responses.

Students were given 5 minutes to review the case and asked to verbally assess and provide recommendations on how they would replace potassium for the patient, including specifics on potassium goal, dose, route, lab frequency, and relevant concurrent interventions. Following student analysis, preceptors mentored student responses by prompting additional responses, filling in student knowledge gaps, and providing alternative views on the hypokalemia vignettes. Sessions concluded with preceptors summarizing the patient case by sharing their overall view and approach to the specific patient case. on the case. Virtual sessions and recordings were held on the Microsoft Teams Platform. All participants signed consent forms for recorded sessions via RedCap^®^.^9^ Consent forms and preceptor instructions for recorded sessions are available in the **Supplemental Appendix**.

### Model, Experimental Conditions, and prompting

GPT-5-Chat was evaluated against the 20 patient cases dataset and prompted to answer 6 questions involving potassium goal, dosing, route of administration, lab frequency, and concurrent interventions. The model was also asked to categorize its perceived level of confidence for responses and complexity of each case. Each of the 6 questions was posed as an independent API call with no shared chat context, and the model was constrained to reply with a single JSON object selecting from a fixed list of multiple-choice options. Two experiment conditions were assessed: the LLMs base prompt performance and performance when provided the investigator-curated potassium dosing criteria and rules. Each case was run in triplicate. GPT-5-Chat was run with its recommended default temperature 1.2. Complete prompt text is located in the **Supplemental Appendix**.

### Outcomes

The primary outcome was appropriate recommendations for each category in comparison to ground truth clinician answers. Secondary outcomes included error rates within each question and Harm Associated with medication Error Classification (HAMEC) scores.^10^ Errors were defined as any outputs that strayed from clinicians’ responses. Outcomes were manually graded by two critical care pharmacists. To account for practice variability for potassium dosing, a range of greater than or less than 20 mEq from the clinician dosing answers were considered to be correct. HAMEC scores were evaluated by the same pharmacists and graded for risk of harm based on LLM errors.

### Statistical Analysis

Descriptive statistics were used to describe results for GPT-5-Chat outputs in comparison to clinician answers and to summarize external clinician survey responses to patient cases and agreement to clinician panel answers. Accuracy was defined as appropriate answers to each question divided by total responses and multiplied by 100% for the GPT-5-Chat experiment. Errors were defined as any LLM output straying from clinician panel answers.

## Results

### Potassium Dosing Criterion and Rules

Seven pharmacists were involved in curating the potassium dosing guidance document. This clinician panel included a diverse group of pharmacists from different levels of care and specialties (cardiology, internal medicine, and critical care). The default rule for potassium dosing was set to “replace 10 mEq of K+ for every 0.1 mEq/L desired increase in serum potassium levels”.

The panel identified clinical scenarios and created dosing criteria for special populations where the default rule would not be appropriate for replacing potassium. Such special populations deemed important by the clinician panel included: those at risk for or experiencing refeeding syndrome, hypomagnesemia, diabetic ketoacidosis (DKA), renal replacement therapy (RRT), targeted temperature management (TTM) or hypothermia, diuretic administration or increased potassium loss, acute kidney injury (AKI), chronic kidney disease (CKD), or no urine output, cardiac patients, and imminent risk of or cardiac arrest due to hypokalemia. The panel reviewed and specified potassium goals, dosing rules, lab frequency, and concurrent interventions for each special situation.

Non-negotiable rules or “Never break” rules comprised recommendations related to safe and appropriate intravenous (IV) administration of potassium products (i.e. “do NOT administer undiluted or as IV push, rate of administration, electrocardiogram (EKG) monitoring, etc.). Cardiac arrest or imminent cardiac arrest patient were the only exception to non-negotiable rules. Negotiable rules comprised guidance on total daily dosing, oral administration, EKG monitoring, route of administration preference based on serum potassium labs, and frequency of labs based on clinical scenarios.

### Potassium Cases Dataset

Two clinical pharmacists created 20 patient vignettes, while five clinical pharmacists reviewed and validated the hypokalemia patient cases for clinical accuracy and agreement with clinician answers. Patient cases were curated to include a mix of clinical scenarios where potassium dosing could follow either the default rule, or one to two of the special scenarios from the potassium dosing guidance document.

### Clinician Survey

A total of 54 surveys were attempted, while only 3 surveys were returned completed. Survey baseline characteristics and potassium case responses can be seen in **Table 1**. Across all cases, the clinicians most commonly labeled their level of confidence and level of complexity for cases as having high confidence and low complexity (39%), or as high confidence and high complexity (30%). Overall agreement with the clinician panel answers for cases weighed the most in “highly agree” and “somewhat agree” category with 28.6% and 38.2%, respectively. Clinicians were more likely to disagree with the potassium goal (22.2%) and route of administration (24.4%) in comparison to the clinical panel answers.

**Table 1.** Clinician survey results.

| Variable | Clinician responses,<br>Cases n = 20 |
| --- | --- |
| *Baseline Characteristics* |  |
| Institution, n (%) |  |
| UCHealth UCH | 27/34 (79.4) |
| Wellstar MCG | 5/34 (14.7) |
| UNC | 2/34 (5.9) |
| Current role |  |
| Pharmacist | 29/52 (55.8) |
| Physician | 11/52 (21.2) |
| Nurse Practitioner | 7/52 (13.5) |
| Physician Assistant | 5/52 (9.6) |
| Specialty |  |
| Ambulatory Care | 4/52 (7.7) |
| Burn | 1/52 (1.9) |
| Cardiology | 5/52 (9.6) |
| Critical Care | 10/52 (19.2) |
| Emergency Medicine | 1/52 (1.9) |
| Hepatology | 6/52 (11.5) |
| Infectious Disease | 1/52 (1.9) |
| Internal/Family Medicine | 17/52 (32.7) |
| Nephrology | 2/52 (3.8) |
| Neurology | 2/52 (3.8) |
| Oncology | 1/52 (1.9) |
| Surgery/Trauma | 2/52 (3.8) |
| Level of care |  |
| Intensive Care Unit | 14/51 (27.5) |
| Intermediate | 18/51 (35.3) |
| Emergency Department | 7/51 (13.7) |
| General Ward | 27/51 (52.9) |
| Outpatient | 3/51 (5.9) |
| Other | 3/51 (5.9) |
| Highest level of training |  |
| Fellowship (complete) | 8/52 (15.4) |
| Fellowship (in process) | 18/52 (34.6) |
| Residency (complete) | 20/52 (38.5) |
| Residency (in process) | 2/52 (3.8) |
| No formal training | 4/52 (7.7) |
| Highest formal training |  |
| PhD | 4/52 (7.7) |
| MD, PharmD, DO, etc. | 41/52 (78.8) |
| Masters | 6/52 (11.5) |
| Bachelors | 0/52 (0) |
| Other | 1/52 (1.9) |
| Years of experience |  |
| 0 or in training | 3/52 (5.8) |
| 1-2 years | 14/52 (26.9) |
| 3-5 years | 7/52 (13.5) |
| 6-10 years | 18/52 (34.6) |
| Over 10 years | 10/52 (19.2) |
| *Case results* |  |
| Confidence vs Complexity, n (%) |  |
| High Confidence, Low Complexity | 39/100 (39.0) |
| High Confidence, High Complexity | 30/100 (30.0) |
| Low Confidence, Low Complexity | 14/100 (14.0) |
| Low Confidence, High Complexity | 17/100 (17.0) |
| Overall Agreement with clinician panel answers, n (%) |  |
| Highly agree | 30/105 (28.6) |
| Somewhat agree | 40/105 (38.2) |
| Neither | 10/105 (9.5) |
| Somewhat disagree | 15/105 (14.3) |
| Highly disagree | 5/105 (4.7) |
| I'm not sure | 5/105 (4.7) |
| Disagree with specific answers, n (%) |  |
| Goal | 20/90 (22.2) |
| Dose | 17/90 (18.9) |
| Route | 22/90 (24.4) |
| Lab frequency | 15/90 (16.7) |
| Concurrent interventions | 5/90 (5.6) |
| NA- agree with all | 11/90 (12.2) |

This table summarizes the data of baseline characteristics and results from the external clinician survey responses. Data for Confidence versus complexity, overall agreement, and disagreement with specific answers are an accumulation of responses across all 20 patient cases.
UCH: University of Colorado Hospital; MCG: Medical College of Georgia; UNC: University of North Carolina; PhD: Doctor of Philosophy; MD: Doctor of Medicine; PharmD: Doctor of Pharmacy; DO: Doctor of Osteopathic Medicine; NA: not applicable; n: sample size.

### Preceptor-student recordings

Four recorded sessions were completed, involving participation from three clinical pharmacist preceptors and six pharmacy students. Pharmacy candidate years of training varied from second-year to fourth year students. In total, eight of the 20 patient cases from the potassium dataset were reviewed.

### Model prompting

Clinicians graded a total of 120 GPT-5-Chat queries—60 with, and 60 without, the potassium replacement criteria. When given the potassium dosing guidance document, total errors dropped from 165 to 104, and average accuracy improved from 45% to 65%, when the criteria was provided. GPT-5-Chat expressed a high level of confidence for 100% of responses, while labeling 80% and 76% of cases as highly complex with and without the criteria, respectively. Error rates range from 43-83% in the no criteria group, versus 10-68% in the criteria group. The highest category of error rates occurred when prompting recommendations on concurrent interventions, followed by potassium dose. Significant HAMEC scores were described in both the “without criteria” and “with criteria” groups. However, HAMEC scores of 2-4 greatly reduced after providing GPT-5-Chat with the potassium dosing guidance document. Complete results are reported in **Table 2**.

**Table 2.** GPT-5-Chat Hypokalemia Case Evaluation.

| Variable | Without dosing criteria | With dosing criteria |
| --- | --- | --- |
| Goal accuracy, n (%) | 27 (45.0) | 54 (90.0) |
| Dose accuracy*, n (%) | 7 (11.7) | 22 (36.7) |
| Dose accuracy ± 20 mEq*, n (%) | 33 (55.0) | 39 (65.0) |
| Route accuracy, n (%) | 31 (51.7) | 34 (56.7) |
| Lab frequency accuracy, n (%) | 33 (55.0) | 42 (70.0) |
| Concurrent interventions accuracy, n (%) | 10 (16.7) | 19 (31.7) |
| Total errors, n | 165 | 104 |
| Confidence vs Complexity, n (%) |  |  |
| High Confidence, Low Complexity | 14 (23.3) | 12 (20.0) |
| High Confidence, High Complexity | 46 (76.7) | 48 (80.0) |
| Low Confidence, Low Complexity | 0 | 0 |
| Low Confidence, High Complexity | 0 | 0 |
| HAMEC Scores, n (%) |  |  |
| 0 | 0 | 2 (3.0) |
| 1 | 23 (38.3) | 42 (70.0) |
| 2 | 17 (28.3) | 9 (15.0) |
| 3 | 17 (28.3) | 7 (11.7) |
| 4 | 3 (5.0) | 0 |

Table 2 demonstrates the increase in accuracy as more dosing criteria is given. The confidence and complexity levels are comparable between the two criteria. Harm Associated with Medication Error Classification (HAMEC) is an identification tool to measure potential or actual patient harm related to a medication event and the degree of how harmful it is. Using HAMEC potential harm classification, 0= no harm, 1= minor, 2= moderate, 3= serious, and 4= severe. HAMEC: Harm Associated with Medication Error Classification; mEq: milliequivalent, n: sample size. Dose accuracy is defined as the rate of exactly correct answers (i.e. the LLM suggested 40mEq as the appropriate repletion dose and the clinician panel/guideline also would recommend 40mEq). Dose accuracy ± 20 mEq was defined as the rate of correct answers that were within 20mEq of the desired answer per the clinician panel (i.e. if the clinician panel/guideline recommended 40mEq, an LLM answer of anywhere between 20mEq and 60mEq would have been deemed correct).

## Discussion

This analysis is the first to our knowledge to create a comprehensive potassium dosing guide for assessment of a large language model performance on complex potassium dosing patient cases. Study findings show that external clinicians tend to, more commonly, agree with the curated potassium dosing criteria and rules, while GPT-5-Chat’s performance also improved with this guidance. However, this improvement is marginal and does not represent safe and effective application of artificial intelligence for decision support of electrolyte replacement. Risk for potential harm was also reduced with no HAMEC category 4 (severe harm) scores in the “with criteria” group compared to 3 instances in the “without criteria” group. Category 3 (serious harm) and category 2 (moderate harm) scores also showed considerably lower in the “with criteria” group. Though an improvement, the frequency of potential harm is unsuitable for implementation in practice and would need to showcase virtually no errors to meet safety benchmarking.

Utilizing machine learning models for administrative and decision-support tools has become a trending subject in healthcare with potential to alleviate workload strain and increase time spent at the bedside.^11-13^ However, treatment tasks related to comprehensive medication management (CMM) continue to be a challenging area for benchmarking across all LLMs. Performance must match or exceed current standard of practice with both safety and efficacy evaluations being the top priority of assessments.^14-21^ For example, in this study evaluation GPT-5-Chat recommended the administration of over 200 mEq of potassium, only by intravenous route and prior to the next laboratory check, for a patient with a severely low serum potassium level of 1.1 mEq/L. Within this specific case, the patient was described to be stable, with no EKG changes and had an acute kidney injury. Though an urgent need for potassium replacement, a dose of greater than 200 mEq of potassium without a laboratory recheck in someone with an acute kidney injury could result in overcorrect and detrimental outcomes for the patient. Such critical thinking and reasoning skill are lacking in current performance of large language models.

This analysis has several limitations. First, the potassium criteria and rules were created and reviewed solely by pharmacists. Although a highly qualified group for potassium dosing, it may not be inclusive of other clinician types (i.e., physician, NP, PA, etc.). Additionally, the clinician survey had poor completion rates. Numbers of completed cases dropped considerably after patient case #6, resulting in other cases being evaluated only by a small group of clinicians and may not capture external validation of the curated potassium dosing criteria and rules or practice variability. GPT-5-Chat was also assessed similarly to the clinician survey, with responses being limited to a list of potential options for each prompted question as opposed to a true free answer structure.

Future work should extend potassium dosing evaluation beyond static criteria and multiple-choice outputs toward dynamic, reasoning-based supervision. Although providing a dosing criteria document improved model accuracy, persistent errors in dose selection and concurrent interventions suggest that models need supervision on how to use criteria in context. The preceptor-student recordings collected in this study provide a natural next step: they capture how pharmacists elicit an initial plan, ask targeted questions, correct missed cues, and guide trainees toward safer potassium recommendations. These dialogues can be annotated into atomic medication facts and pharmacist-derived reasoning rubrics, enabling future benchmarks to evaluate not only whether a model selects the correct potassium goal, dose, route, lab frequency, or concurrent intervention, but also whether it recognizes the patient-specific cues and safety constraints that justify those choices.^22^

## Conclusion

Benchmarks must appropriately reflect clinical complexity to be considered valuable for the deployment of agentic artificial intelligence tools in the healthcare domain. While LLMs appeared capable of following a single dosing rule, substantial safety concerns arose when patient and task complexity increased, and GPT-5-Chat recommended potassium repletion that had potential for patient harm.

**Figure 1.**
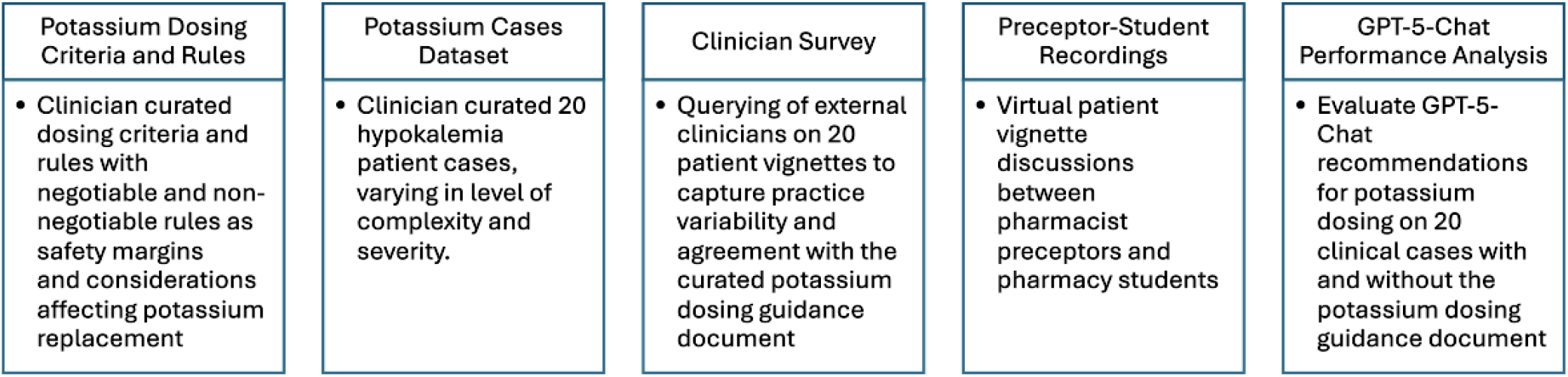
Methodologic Framework

## Data Availability

All data produced in the present work are contained in the manuscript

## Notes

**Conflicts of Interest:** The authors have no conflicts of interest.

### Competing Interest Statement

The authors have declared no competing interest.

## References

1. Hodkinson A, Tyler N, Ashcroft DM, et al. Preventable medication harm across health care settings: a systematic review and meta-analysis. BMC Med. Nov 6 2020;18(1):313. doi:10.1186/s12916-020-01774-9

2. Chase AM, Azimi HA, Forehand CC, et al. An Evaluation of the Relationship Between Medication Regimen Complexity as Measured by the MRC-ICU to Medication Errors in Critically Ill Patients. Hosp Pharm. Dec 2023;58(6):569–574. doi:10.1177/00185787231170386

3. Practices IfSM. About Us. Accessed May 28, 2026. https://home.ecri.org/pages/ismp-about-us

4. Practices IfSM. ISMP List of High-Alert Medications in Acute Care Settings. Accessed May 28, 2026. https://www.ismp.org/system/files/resources/2024-01/ISMP_HighAlert_AcuteCare_List_010924_MS5760.pdf

5. Kraft MD, Btaiche IF, Sacks GS, Kudsk KA. Treatment of electrolyte disorders in adult patients in the intensive care unit. Am J Health Syst Pharm. Aug 15 2005;62(16):1663–82. doi:10.2146/ajhp040300

6. Jiang Y, Black KC, Geng G, et al. MedAgentBench: A Virtual EHR Environment to Benchmark Medical LLM Agents. NEJM AI. 2025;2(9):AIdbp2500144. doi:doi:10.1056/AIdbp2500144

7. Association WM. World Medical Association Declaration of Helsinki: Ethical Principles for Medical Research Involving Human Subjects. JAMA. 2013;310(20):2191–2194. doi:10.1001/jama.2013.281053

8. Gallifant J, Afshar M, Ameen S, et al. The TRIPOD-LLM reporting guideline for studies using large language models. Nat Med. Jan 2025;31(1):60–69. doi:10.1038/s41591-024-03425-5

9. Harris PA, Taylor R, Minor BL, et al. The REDCap consortium: Building an international community of software platform partners. J Biomed Inform. Jul 2019;95:103208. doi:10.1016/j.jbi.2019.103208

10. Gates PJ, Baysari MT, Mumford V, Raban MZ, Westbrook JI. Standardising the Classification of Harm Associated with Medication Errors: The Harm Associated with Medication Error Classification (HAMEC). Drug Saf. Aug 2019;42(8):931–939. doi:10.1007/s40264-019-00823-4

11. Meskó B, Görög M. A short guide for medical professionals in the era of artificial intelligence. NPJ Digit Med. 2020;3:126. doi:10.1038/s41746-020-00333-z

12. Pavuluri S, Sangal R, Sather J, Taylor RA. Balancing act: the complex role of artificial intelligence in addressing burnout and healthcare workforce dynamics. BMJ Health Care Inform. Aug 24 2024;31(1)doi:10.1136/bmjhci-2024-101120

13. Maleki Varnosfaderani S, Forouzanfar M. The Role of AI in Hospitals and Clinics: Transforming Healthcare in the 21st Century. Bioengineering (Basel). Mar 29 2024;11(4)doi:10.3390/bioengineering11040337

14. Liu Z, Xu S, Wu Z, et al. PharmacyGPT: exploration of artificial intelligence for medication management in the intensive care unit. BMC Med Inform Decis Mak. Oct 28 2025;25(1):398. doi:10.1186/s12911-025-03230-1

15. Chase A, Most A, Xu S, et al. Large language models management of complex medication regimens: a case-based evaluation. Front Pharmacol. 2025;16:1514445. doi:10.3389/fphar.2025.1514445

16. Gilbert S, Harvey H, Melvin T, Vollebregt E, Wicks P. Large language model AI chatbots require approval as medical devices. Nat Med. Oct 2023;29(10):2396–2398. doi:10.1038/s41591-023-02412-6

17. Yang H, Hu M, Most A, et al. Evaluating accuracy and reproducibility of large language model performance on critical care assessments in pharmacy education. Front Artif Intell. 2024;7:1514896. doi:10.3389/frai.2024.1514896

18. Blotske K, Zhao X, Henry K, et al. Drug-drug interaction identification using large language models. medRxiv. Dec 29 2025;doi:10.64898/2025.12.03.25341549

19. Henry K, Smith B, Zhao X, et al. Drug or Pokémon? An analysis of the ability of large language models to discern fabricated medications. medRxiv. Jan 13 2026;doi:10.64898/2026.01.12.26343930

20. Blotske K, Zhao X, Cargile M, et al. MedMatch: a first step for the automation of large language model performance benchmarking for medication-related tasks. medRxiv. Jan 15 2026;doi:10.64898/2026.01.13.26343949

21. Zhao X, Blotske K, Cargile M, et al. Rx-LLM: a benchmarking suite to evaluate safe large language model performance for medication-related tasks. medRxiv. Dec 30 2025;doi:10.64898/2025.12.01.25341004

22. Gong L, Fang W, Yang T, et al. MedDialogRubrics: A Comprehensive Benchmark and Evaluation Framework for Multi-turn Medical Consultations in Large Language Models. 2026.

