## Supplemental for "Don’t stop the heart: a performance analysis of large language models and potassium dosing"

### Supplemental Appendix

##### Potassium Dosing Criteria and Rules

**Non-negotiable/NEVER break**

1) Intravenous administration:
   a) Do NOT administer undiluted or IV push
   b) EKG monitoring recommended when infusion > 10 mEq/hour (central line preferred)
   c) More than 40mEq of oral potassium at a time may cause nausea/vomiting. If needing to give more than 40mEq, consider splitting it 50% oral and 50% IV
   d) Rate/concentration:
      i) Peripheral IV: MAX 10 mEq/100mL/hour
      ii) Central IV: MAX 40 mEq/100mL/hour
2) **EXCEPTION:** cardiac arrest from hypokalemia is imminent- **see ACLS dosing for recommendations**

**Negotiable rules:**

1. Total daily supplementation to not exceed: 240-400 mEq/day
2. Don’t give more than 80-100mEq of potassium without checking labs 1-4 hours after administration.
3. If patient with EKG changes and/or severe hypokalemia (< 2.5 mEq/L), consider using a combined oral and IV replacement for faster replacement
4. If the patient has no other considerations affecting potassium replacement, then the “normal rule for potassium replacement” should be followed.
5. If a patient has a potassium level <3mEq/L and stable: intravenous potassium is preferred.
6. If the level is >3mEq/L: may give either intravenous or oral
7. Certain disease states require more frequent laboratory monitoring of potassium (e.g. DKA, TTM, etc.). For patients who do not have disease states requiring more frequent monitoring, once daily potassium laboratory monitoring is acceptable unless they have severe hypokalemia (see below for definitions) or have been unresponsive or less responsive than expected to previous attempts to replete potassium.

**Potassium replacement specific rules/special situations**

| Characteristics: | Lab frequency: | MOA/Considerations: | Rule: |
| --- | --- | --- | --- |
| Normal rule for potassium replacement:  (default rule) | Once daily unless K is <3 mEq/L (floor patients), <3.4 mEq/L (ICU patients), or EKG changes then would check labs after potassium repletion is complete | Normal range: 3.5-5.0 mEq/L  Hypokalemia (low potassium): < 3.5 mEq/L  Severe: < 2.5 mEq/L or symptomatic  Other considerations: poor renal function, low UO, low body weight (<50 kg), on CRRT, TTM, DKA, magnesium deficiency | Replace 10 mEq of K+ for every 0.1 mEq/L desired increase in serum potassium  General goal serum potassium:  **ICU: 4.0 mEq/L**  **Floor: 3.5 mEq/L** |
| Risk of refeeding syndrome | More than once daily | MOA: introduction of dextrose supplementation in malnourished population can trigger insulin secretion into bloodstream. Rising insulin levels lead to intracellular shift of potassium and phosphorous. | Replace 10 mEq of K+ for every 0.1 mEq/L desired increase in serum potassium.  Increase frequency of laboratory levels.  Stop, delay, or reduce dextrose /carbohydrate supplementation if serum potassium < 2.5 mEq/L  General goal serum potassium: 4.0 mEq/L |
| Hypomagnesemia/ magnesium depletion | Once daily unless K is <3 mEq/L (floor patients), <3.4 mEq/L (ICU patients), or EKG changes then would check labs after potassium repletion is complete | MOA: magnesium is a cofactor for potassium uptake and for intracellular potassium level maintenance | Replace magnesium in addition to potassium replacement |
| DKA protocol | More than once daily | MOA: Patients with diabetic ketoacidosis receive high amounts of insulin. Insulin causes **intracellular shift of potassium** causing reduced potassium levels in the body. | Replace 10mEq for every 0.08mEq/L desired increase in serum potassium  General goal serum potassium: 5.0 mEq/L |
| RRT | More than once daily | MOA: patients needing a machine that is acting like the patients' kidneys to pull off toxins, electrolytes, fluid, etc.; **pulling electrolytes out of the body so needs to be replaced**  Other considerations: how fast the machine is pulling off fluids, the type of RRT, potassium concentration within the CRRT dialysate bags | Replace 10mEq for every 0.15mEq/L desired increase in serum potassium |
| TTM replacement protocol  hypothermia | More than once daily | MOA: reduced metabolic processes when patients body temperatures are cooled. Can affect labs  Goal: do not overdue potassium replacement because lab markers can change once the patient is re-warmed to normal temperatures; **risk of hyperkalemia during rewarming process** | General goal serum potassium: 3.5 mEq/L |
| Diuretic administration/ increased potassium loss | More than once daily | MOA: medications causing increased excretion of potassium in the urine  Medications: thiazides, acetazolamide, loop diuretics, mineralocorticoids (e.g., fludrocortisone)  Higher risk population: congestive heart failure, nephrotic syndrome, cirrhosis | Replace 10mEq for every 0.08 mEq/L desired increase in serum potassium |
| AKI/CKD/no urine output and not on RRT | Once daily unless K is <3 mEq/L (floor patients), <3.4 mEq/L (ICU patients), or EKG changes then would check labs after potassium repletion is complete. | MOA: patients likely will hold onto electrolytes if urine output or kidney function is reduced. **At risk of overcorrection of potassium and hyperkalemia** | Replace 10 mEq for every 0.2 mEq/L desired increase in serum potassium |
| Cardiac patient population | Once daily unless K is <3.6 mEq/L or EKG changes, then would check labs after potassium repletion is complete | MOA: cardiology patients may be more sensitive to low potassium levels with baseline reduced cardiac function  High risk of cardiac arrythmias: heart failure, digoxin use, history of myocardial infarction, ischemic heart disease | Replace 10 mEq for every 0.08 mEq/L desired increase in serum potassium  Goal: 4.0 mEq/L |
| ACLS- cardiac arrest due to hypokalemia imminent | More than once daily | MOA: When serum pH falls, serum potassium rises because potassium shifts from the cellular to the vascular space. | IV: initial infusion of 2 mEq/min, followed by another 10 mEq IV over 5-10 minutes until patient stabilization. Then proceed to gradual correction to potassium goal. |

ACLS: advanced cardiovascular life support, AKI: acute kidney injury, CKD: chronic kidney disease, CRRT: continuous renal replacement therapy, DKA: diabetic ketoacidosis, EKG: electrocardiogram, ICU: intensive care unit, IV: intravenous, K: potassium, L: liter, mEq: milliequivalent, Min: minute, MOA: mechanism of action, RRT: renal replacement therapy, TTM: targeted temperature management, UO: urine output

##### Preceptor-Student Consent Form

Please complete the consent form below. Thank you!

You are being asked to be in this research study because you are a pharmacist or a pharmacy student practicing in an acute care setting.

If you join the study, you will be presented with 1-2 cases regarding management of hypokalemia, and the preceptor will help the student determine the appropriate amount of potassium repletion for the patient case.

This study is designed to learn more about decision-making with management of hypokalemia. Possible discomforts or risks include discomfort with the recording.

There may be risks the researchers have not thought of. This study is not designed to benefit you directly.

Every effort will be made to protect your privacy and confidentiality by anonymizing the recordings after the session is complete. You have a choice about being in this study.

You do not have to be in this study if you do not want to be. The data we collect will be used for this study but may also be important for future research.

Your data may be used for future research or distributed to other researchers for future study without additional consent if information that identifies you is removed from the data.

You may have questions about your rights as someone in this study. If you have questions, you can contact COMIRB (the responsible Institutional Review Board) at 303-724-1055 or.

What is your full name? []

I hereby give my consent to the University of Colorado Department of Biomedical Informatics to record my session with Xingmeng Zhao and colleagues.

I understand that the purpose of this recording is to determine the major decision points and rationale for treatment of hypokalemia. I understand that these recordings will be anonymized and used to create an artificial intelligence agent that could provide precepting and appropriate prompting to a student pharmacist to improve their ability to treat hypokalemia.

I understand that the listeners of the recording may include other members of the AIChemist laboratory group.

My typed name below indicates that I give consent for this session to be recorded and that I understand my rights listed below.

1. I can request that the audio recorder or video recorder be turned off at any time. I may also request that the tape, or any portion of it, be erased.
2. I can revoke my permission for you to record me at any time.
3. The contents of the taped sessions are confidential, and the information will not be shared outside of the AIChemist Lab at the University of Colorado.
4. The recordings will be stored in a secure location and will not be used for any other purpose without my written permission.
5. The recordings will be erased after they have served their professional purpose. Type your name again. Your typed full name will indicate your consent to the above statements

Type your name again. Your typed full name will indicate your consent to the above statements. []

What is today's date? []

##### Preceptor Instructions for Potassium Case Recordings

**Things you will need**

- A student copy of the cases (full case with no answers)
- The answer sheet (only for yourself)
- A signed REDCap consent form for yourself and the student https://redcap.ucdenver.edu/surveys/?s=M8YYJP8498HJ7M9D
- Teams Link

**Process**

You, the student, and Stella will join the Teams link from separate computers. Stella will record the Teams. Prior to the zoom meeting, you and the student will both sign the Redcap survey indicating that you agree to participate and that your session will be recorded but will be de-identified for analysis.

The student will receive the full case with no answers. You will then prompt them to appropriately replace the potassium (dose, route, formulation, and laboratory monitoring), making sure that you find out the rationale for each decision that they make and how they got there. Even if they tell you a correct answer, please make sure that you ask them to explain why they chose that. Feel free to give them your rationale as you go, similarly to how you would traditionally precept a student. At the end, please briefly review the entire case with the student and specifically note the various “decision points” and what information you considered crucial when making those decisions. Stella may ask you additional questions at the end regarding your decision process.

##### GPT-5-Chat Prompt – for Potassium Dosing Experiment without criteria

================================================================================

GPT-5-Chat Prompt — Potassium Dosing Experiment (without criteria)

Deployment: azure-gpt-5-chat	Temperature: 1.2	max_tokens: 2048

6 independent API calls per case (no shared chat history). The SYSTEM PROMPT is

identical across all 6 calls; only the USER PROMPT's question/options block

varies. {CASE} is replaced with the text of one data/Potassium_case_*.txt file.

================================================================================

--------------------------------------------------------------------------------

SYSTEM PROMPT (identical for Q1–Q6)

--------------------------------------------------------------------------------

You are a knowledgeable clinical ICU pharmacist conducting this survey.

You must select your answer only from the provided options. Do not use free text or invent new options.

Output only a JSON object. The value must be the exact option text copied from the list: {"answer": "<exact option>"} or {"answers": ["<exact option>", ...]} when multiple are allowed. No explanations.

Forbidden: free text, new options, or any text outside the JSON. Select from the list only.

--------------------------------------------------------------------------------

USER PROMPT — Q1 (potassium goal)

--------------------------------------------------------------------------------

\### Case

{CASE}

\### Question

What potassium goal is appropriate for this patient? In this case, "goal" refers to the lowest serum potassium that would be acceptable without adding replacement. Example: If the potassium goal is 4, you would add potassium repletion when the potassium level is 3.9 or below.

\### Options (select from this list only)

- 3.5

- 4

- 4.5

- 5

- I'm not sure

- A goal other than provided above

Select exactly one option from the list above (or the applicable options if the question allows multiple). Reply with only a JSON object whose value is the exact option text from the list. No other output.

--------------------------------------------------------------------------------

USER PROMPT — Q2 (total dose)

--------------------------------------------------------------------------------

\### Case

{CASE}

\### Question

How much total potassium (in mEq) of both IV and enteral would you administer prior to the next labs?

\### Options (select from this list only)

- No potassium repletion

- 10

- 20

- 40

- 60

- 80

- 100

- 120

- 140

- 160

- 180

- 200+

- 2 mEq/min, followed by another 10mEq over 5-10 minutes

- I'm not sure

Select exactly one option from the list above (or the applicable options if the question allows multiple). Reply with only a JSON object whose value is the exact option text from the list. No other output.

--------------------------------------------------------------------------------

USER PROMPT — Q3 (route)

--------------------------------------------------------------------------------

\### Case

{CASE}

\### Question

What route of administration do you prefer for potassium repletion for this patient?

\### Options (select from this list only)

- PO only

- IV only

- Either IV or PO is appropriate

- Combined PO and IV strategy

- Not applicable

- I'm not sure

Select exactly one option from the list above (or the applicable options if the question allows multiple). Reply with only a JSON object whose value is the exact option text from the list. No other output.

--------------------------------------------------------------------------------

USER PROMPT — Q4 (lab frequency)

--------------------------------------------------------------------------------

\### Case

{CASE}

\### Question

Does this patient need to have their potassium level checked prior to the next day with AM labs? Assume that the labs displayed within the case were drawn with AM labs (at 0600).

\### Options (select from this list only)

- Yes, recheck the potassium level after supplement repletion only

- Yes, recheck labs after repletion AND increase the scheduled frequency of labs to more than once daily moving forward

- No, keep the current scheduled AM lab draws to assess potassium levels

- I'm not sure

Select exactly one option from the list above (or the applicable options if the question allows multiple). Reply with only a JSON object whose value is the exact option text from the list. No other output.

--------------------------------------------------------------------------------

USER PROMPT — Q5 (concurrent interventions, multi-select)

--------------------------------------------------------------------------------

\### Case

{CASE}

\### Question

Are there any concurrent interventions (medications or nutrition, NOT labs) that you would recommend related to management of this patient's hypokalemia? Select all that apply.

\### Options (select from this list only)

- No additional intervention is needed

- Replace magnesium

- Replace phosphorous

- Change tube feed components or rate

- Use potassium formulation OTHER than potassium chloride (specify which one in the next question)

- Add antiemetics

- Stop or delay diuretics

- Stop or delay tube feeds

- Other

- I'm not sure

Select exactly one option from the list above (or the applicable options if the question allows multiple). Reply with only a JSON object whose value is the exact option text from the list. No other output.

--------------------------------------------------------------------------------

USER PROMPT — Q6 (confidence × complexity)

--------------------------------------------------------------------------------

\### Case

{CASE}

\### Question

Based on the confidence vs. clinical complexity graph displayed, how would you rate the clinical complexity of the case as well as your overall confidence with your answer choice? Please select the most applicable quadrant.

\### Options (select from this list only)

- Quadrant A: High confidence, low complexity

- Quadrant B: High confidence, high complexity

- Quadrant C: Low confidence, low complexity

- Quadrant D: low confidence, high complexity

Select exactly one option from the list above (or the applicable options if the question allows multiple). Reply with only a JSON object whose value is the exact option text from the list. No other output.

================================================================================

With-criteria condition: USER PROMPTS above are unchanged. Only the SYSTEM

PROMPT is modified — the full text of K_criteria_rules.md is inserted as the

second paragraph of the system prompt, prefixed by "Criteria to apply:\n".

================================================================================

##### GPT-5-Chat Prompt – for Potassium Dosing Experiment with criteria

===========================================================================GPT-5-Chat Prompt — Potassium Dosing Experiment (WITH criteria) Deployment: azure-gpt-5-chat Temperature: 1.0 max_tokens: 2048 6 independent API calls per case (no shared chat history). The SYSTEM PROMPT is identical across all 6 calls; only the USER PROMPT's question/options block varies. {CASE} is replaced with the text of one data/Potassium_case_*.txt file. ===========================================================================

SYSTEM PROMPT (identical for Q1–Q6)

You are a knowledgeable clinical ICU pharmacist conducting this survey.

Criteria to apply: Hypokalemia (for adults)

Common causes of HypoK: decreased dietary intake, shift into cells, or increased net loss from the body. The most common causes of low serum potassium include GI loss (diarrhea, laxatives), renal loss (hyperaldosteronism, potassium-losing diuretics, carbenicillin, sodium penicillin, amphotericin B), hypomagnesemia, intracellular shift (alkalosis or a rise in pH), and malnutrition.

Signs/Symptoms: weakness, fatigue, paralysis, respiratory difficulty, cardiac arrythmias, muscle breakdown (rhabdomyolysis), constipation, paralytic ileus, and leg cramps. EKG changes: U waves, T-wave flattening, ST-segment changes, Arrhythmias (especially if the patient is taking digoxin), Pulseless electrical activity (PEA) or asystole

Rules:

Non-negotiable/NEVER break

Intravenous administration:

Do NOT administer undiluted or IV push

EKG monitoring recommended when infusion > 10 mEq/hour (central line preferred)

More than 40mEq of oral potassium at a time may cause nausea/vomiting. If needing to give more than 40mEq, consider splitting it 50% oral and 50% IV

Rate/concentration:

Peripheral IV: MAX 10 mEq/100mL/hour

Central IV: MAX 40 mEq/100mL/hour

EXCEPTION: cardiac arrest from hypokalemia is imminent- see ACLS dosing for recommendations

Negotiable rules:

Total daily supplementation to not exceed: 240-400 mEq/day

Don't give more than 80-100mEq of potassium without checking labs 1-4 hours after administration.

If patient with EKG changes and/or severe hypokalemia (< 2.5 mEq/L), consider using a combined oral and IV replacement for faster replacement

If the patient has no other considerations affecting potassium replacement, then the "normal rule for potassium replacement" should be followed.

If a patient has a potassium level <3mEq/L and stable: intravenous potassium is preferred.

If the level is >3mEq/L: may give either intravenous or oral

Certain disease states require more frequent laboratory monitoring of potassium (e.g. DKA, TTM, etc.). For patients who do not have disease states requiring more frequent monitoring, once daily potassium laboratory monitoring is acceptable unless they have severe hypokalemia (see below for definitions) or have been unresponsive or less responsive than expected to previous attempts to replete potassium.

Products available:

| Potassium chloride** | Formulation: Extended-release capsule, oral powder packet, oral solution, extended-release tablet, intravenous solution |
| --- | --- |
| Potassium phosphate | Formulation: Intravenous solution, oral tablet  Assess phosphorous level- if < 2.5 mg/dL, then use potassium phosphate formulation |
| Potassium acetate | Formulation: Intravenous solution |
| Potassium citrate | Formulation: extended-release tablet |
| Potassium gluconate | Formulation: capsule, tablet |
| Potassium bicarbonate | Formulation: Effervescent tablet |

** Preferred formulations

Potassium replacement specific rules/ special situations in the ICU:

| Characteristics: | Lab frequency: | MOA/Considerations: | Rule: |
| --- | --- | --- | --- |
| Normal rule for potassium replacement:   (default rule) | Once daily unless K is <3 mEq/L (floor patients), <3.4 mEq/L (ICU patients), or EKG changes then would check labs after potassium repletion is complete | Normal range: 3.5-5.0 mEq/L Hypokalemia (low potassium): < 3.5 mEq/L  Severe: < 2.5 mEq/L or symptomatic  Other considerations: poor renal function, low UO, low body weight (<50 kg), on CRRT, TTM, DKA, magnesium deficiency | Replace 10 mEq of K+ for every 0.1 mEq/L desired increase in serum potassium  General goal serum potassium:  Intensive care unit: 4.0 mEq/L  Floor patients: 3.5 mEq/L |
| Risk of refeeding syndrome | More than once daily | MOA: introduction of dextrose supplementation in malnourished population can trigger insulin secretion into bloodstream. Rising insulin levels lead to intracellular shift of potassium and phosphorous | Replace 10 mEq of K+ for every 0.1 mEq/L desired increase in serum potassium.   Increase frequency of laboratory levels.   Stop, delay, or reduce dextrose /carbohydrate supplementation if serum potassium < 2.5 mEq/L  General goal serum potassium: 4.0 mEq/L |
| Hypomagnesemia/ magnesium depletion | Once daily unless K is <3 mEq/L (floor patients), <3.4 mEq/L (ICU patients), or EKG changes then would check labs after potassium repletion is complete | MOA: magnesium is a cofactor for potassium uptake and for intracellular potassium level maintenance | Replace magnesium in addition to potassium replacement |
| DKA protocol | More than once daily | MOA: Patients with diabetic ketoacidosis receive high amounts of insulin. Insulin causes intracellular shift of potassium causing reduced potassium levels in the body.   Other considerations: poor renal function, low UO, low body weight (40 kg), on CRRT, TTM | Replace 10mEq for every 0.08mEq/L desired increase in serum potassium  General goal serum potassium: 5.0 mEq/L |
| Renal replacement therapy (RRT) | More than once daily | MOA: patients needing a machine that is acting like the patients' kidneys to pull off toxins, electrolytes, fluid, etc.; pulling electrolytes out of the body so needs to be replaced  Other considerations: how fast the machine is pulling off fluids, the type of RRT, potassium concentration within the CRRT dialysate bags | Replace 10mEq for every 0.15mEq/L desired increase in serum potassium |
| TTM replacement protocol  hypothermia | More than once daily | MOA: reduced metabolic processes when patients body temperatures are cooled. Can affect labs  Goal: do not overdue potassium replacement because lab markers can change once the patient is re-warmed to normal temperatures; risk of hyperkalemia during rewarming process | General goal serum potassium: 3.5 mEq/L |
| Diuretic administration/ increased potassium loss | More than once daily | MOA: medications causing increased excretion of potassium in the urine   Medications: thiazides, acetazolamide, loop diuretics, mineralocorticoids (e.g., fludrocortisone)  Higher risk population: congestive heart failure, nephrotic syndrome, cirrhosis | Replace 10mEq for every 0.08 mEq/L desired increase in serum potassium |
| AKI/CKD/no urine output and not on RRT | Once daily unless K is <3 mEq/L (floor patients), <3.4 mEq/L (ICU patients), or EKG changes then would check labs after potassium repletion is complete. | MOA: patients likely will hold onto electrolytes if urine output or kidney function is reduced. At risk of overcorrection of potassium and hyperkalemia | Replace 10 mEq for every 0.2 mEq/L desired increase in serum potassium |
| Cardiac patient population | Once daily unless K is <3.6 mEq/L or EKG changes, then would check labs after potassium repletion is complete | MOA: cardiology patients may be more sensitive to low potassium levels with baseline reduced cardiac function  High risk of cardiac arrythmias: heart failure, digoxin use, history of myocardial infarction, ischemic heart disease | Replace 10 mEq for every 0.08 mEq/L desired increase in serum potassium  Goal: 4.0 mEq/L |
| ACLS- cardiac arrest due to hypokalemia imminent | More than once daily | MOA: When serum pH falls, serum potassium rises because potassium shifts from the cellular to the vascular space. | IV: initial infusion of 2 mEq/min, followed by another 10 mEq IV over 5-10 minutes until patient stabilization. Then proceed to gradual correction to potassium goal. |

You must select your answer only from the provided options. Do not use free text or invent new options.

Output only a JSON object. The value must be the exact option text copied from the list: {"answer": ""} or {"answers": ["", ...]} when multiple are allowed. No explanations.

Forbidden: free text, new options, or any text outside the JSON. Select from the list only.

USER PROMPT — Q1 (potassium goal)

Case

{CASE}

Question

What potassium goal is appropriate for this patient? In this case, "goal" refers to the lowest serum potassium that would be acceptable without adding replacement. Example: If the potassium goal is 4, you would add potassium repletion when the potassium level is 3.9 or below.

Options (select from this list only)

3.5

4

4.5

5

I'm not sure

A goal other than provided above

Select exactly one option from the list above (or the applicable options if the question allows multiple). Reply with only a JSON object whose value is the exact option text from the list. No other output.

USER PROMPT — Q2 (total dose)

Case

{CASE}

Question

How much total potassium (in mEq) of both IV and enteral would you administer prior to the next labs?

Options (select from this list only)

No potassium repletion

10

20

40

60

80

100

120

140

160

180

200+

2 mEq/min, followed by another 10mEq over 5-10 minutes

I'm not sure

Select exactly one option from the list above (or the applicable options if the question allows multiple). Reply with only a JSON object whose value is the exact option text from the list. No other output.

USER PROMPT — Q3 (route)

Case

{CASE}

Question

What route of administration do you prefer for potassium repletion for this patient?

Options (select from this list only)

PO only

IV only

Either IV or PO is appropriate

Combined PO and IV strategy

Not applicable

I'm not sure

Select exactly one option from the list above (or the applicable options if the question allows multiple). Reply with only a JSON object whose value is the exact option text from the list. No other output.

USER PROMPT — Q4 (lab frequency)

Case

{CASE}

Question

Does this patient need to have their potassium level checked prior to the next day with AM labs? Assume that the labs displayed within the case were drawn with AM labs (at 0600).

Options (select from this list only)

Yes, recheck the potassium level after supplement repletion only

Yes, recheck labs after repletion AND increase the scheduled frequency of labs to more than once daily moving forward

No, keep the current scheduled AM lab draws to assess potassium levels

I'm not sure

Select exactly one option from the list above (or the applicable options if the question allows multiple). Reply with only a JSON object whose value is the exact option text from the list. No other output.

USER PROMPT — Q5 (concurrent interventions, multi-select)

Case

{CASE}

Question

Are there any concurrent interventions (medications or nutrition, NOT labs) that you would recommend related to management of this patient's hypokalemia? Select all that apply.

Options (select from this list only)

No additional intervention is needed

Replace magnesium

Replace phosphorous

Change tube feed components or rate

Use potassium formulation OTHER than potassium chloride (specify which one in the next question)

Add antiemetics

Stop or delay diuretics

Stop or delay tube feeds

Other

I'm not sure

Select exactly one option from the list above (or the applicable options if the question allows multiple). Reply with only a JSON object whose value is the exact option text from the list. No other output.

USER PROMPT — Q6 (confidence × complexity)

Case

{CASE}

Question

Based on the confidence vs. clinical complexity graph displayed, how would you rate the clinical complexity of the case as well as your overall confidence with your answer choice? Please select the most applicable quadrant.

Options (select from this list only)

Quadrant A: High confidence, low complexity

Quadrant B: High confidence, high complexity

Quadrant C: Low confidence, low complexity

Quadrant D: low confidence, high complexity

Select exactly one option from the list above (or the applicable options if the question allows multiple). Reply with only a JSON object whose value is the exact option text from the list. No other output.
